# Rapid and Sensitive Detection of SARS-CoV-2 Infection Using Quantitative Peptide Enrichment LC-MS Analysis

**DOI:** 10.1101/2021.06.02.21258097

**Authors:** Andreas Hober, Tran-Minh Khue Hua, Dominic Foley, Thomas McDonald, Johannes P.C. Vissers, Rebecca Pattison, Samantha Ferries, Sigurd Hermansson, Ingvar Betner, Mathias Uhlen, Morteza Razavi, Richard Yip, Matthew E. Pope, Terry W. Pearson, N. Leigh Anderson, Amy Bartlett, Lisa Calton, Jessica J. Alm, Lars Engstrand, Fredrik Edfors

**Affiliations:** Science for Life Laboratory, Solna, Sweden; The Royal Institute of Technology, Division of Systems Biology, Department of Protein Science, School of Chemistry, Biotechnology and Health, Stockholm. Sweden; Waters Corporation, Wilmslow, UK; Milford, MA; Stockholm, Sweden; SISCAPA Assay Technologies, Inc., Washington DC; Victoria BC Canada; Karolinska Institutet, Department of Microbiology, Tumor and Cell Biology & National Pandemic Center, Karolinska Institutet, Solna, Sweden

**Keywords:** LC-MS, MRM, SARS-CoV-2, NCAP, RT-PCR, digest, peptide, SISCAPA, affinity

## Abstract

Reliable, robust, large-scale molecular testing for SARS-CoV-2 is essential for monitoring the ongoing Covid-19 pandemic. We have developed a scalable analytical approach to detect viral proteins based on peptide immunoaffinity enrichment combined with liquid chromatography - mass spectrometry (LC-MS). This is a multiplexed strategy, based on targeted proteomics analysis and read-out by LC-MS, capable of precisely quantifying and confirming the presence of SARS-CoV-2 in PBS swab media from combined throat/nasopharynx/saliva samples.

The results reveal that the levels of SARS-CoV-2 measured by LC-MS correlate well with their corresponding RT-PCR readout (r=0.79). The analytical workflow shows similar turnaround times as regular RT-PCR instrumentation with a quantitative readout of viral proteins corresponding to cycle thresholds (Ct) equivalents ranging from 21 to 34. Using RT-PCR as a reference, we demonstrate that the LC-MS-based method has 100% negative percent agreement (estimated specificity) and 95% positive percent agreement (estimated sensitivity) when analyzing clinical samples collected from asymptomatic individuals with a Ct within the limit of detection of the mass spectrometer (Ct ≤30). These results suggest that a scalable analytical method based on LC-MS has a place in future pandemic preparedness centers to complement current virus detection technologies.

## Introduction

The severe acute respiratory syndrome coronavirus 2 (SARS-CoV-2) (Wu et al., 2020), leading to the coronavirus disease 2019 (Covid-19), has had a significant impact on human health globally, with more than 234 million confirmed cases (Dong, Du, & Gardner, 2020), assessed October 4th, 2021. The effects of the pandemic are devastating and have led to lockdowns of urban areas across the globe as a response to contain any potential outbreaks (Hale et al., 2021). To monitor the disease, huge investments have been directed towards infrastructure for large-scale testing for ongoing Covid-19 infection (Baker, Wilson, & Anglemyer, 2020). Population-wide screening or cohort testing in the vicinity of an outbreak epicenter is an essential pillar in the global fight against Covid-19 and an indispensable contribution to currently ongoing vaccination programs that pave the way for re-opening societies when entering the endemic phase. Thus, specific molecular diagnostic tools suitable for efficient disease monitoring will play a key role when countries slowly lift their bans on public gatherings, events, and global travel.

The diagnostic method called Real-Time - Polymerase Chain Reaction (RT-PCR) (Freeman, Walker, & Vrana, 1999) is the most widely used technology for detecting SARS-CoV-2 and was established within days after the virus genome was released (Corman et al., 2020). The method is considered as the gold standard by WHO for diagnosing patients with Covid-19 in routine clinical practice. Large-scale laboratories dedicated to PCR-based diagnostics rapidly mobilized worldwide in the early phase of the pandemic, which led to a sudden global shortage of diagnostic reagents (Woolston, 2021). The PCR tests generally have high analytical sensitivity and specificity, even for self-collected samples, often in the range of 95-100% (Altamirano et al., 2020) when evaluated in clinical settings. The observed variance between tests can be partly explained by the inherent sensitivity of the PCR reaction itself or by pre-analytical biases (Lippi, Simundic, & Plebani, 2020), which could lead to either false positive (FP) or false-negative (FN) results. For example, the viral genes can be amplified to detect the virus within days of infection, but the high sensitivity has also been subjected to criticism since it can detect genetic material in circulation not only days after but also multiple weeks after the first day of symptom onset (Lan et al., 2020). The current level of the clinical false-positive rate (FPR) associated with PCR tests is unknown but is dependent on what type of PCR kit and criteria have been used. Some studies report that it can be as much as 4% at certain test facilities (Surkova, Nikolayevskyy, & Drobniewski, 2020). This type of error has the potential to cause the most harm in a scenario entering post Covid-19 when large-volume screening is performed in communities with low prevalence (Healy, Khan, Metezai, Blyth, & Asad, 2021).

As a response to the global shortage, rapid antigen tests have been deployed that directly detect viral antigens. These rapid tests show similar specificity to PCR-based assays (Weissleder, Lee, Ko, & Pittet, 2020), but several studies have shown that they lack sufficient sensitivity when compared to RT-PCR (Fitzpatrick, Pandey, Wells, Sah, & Galvani, 2021; Perchetti, Huang, Mills, Jerome, & Greninger, 2021). Antigen tests also require affinity reagents, an initial bottleneck and a significant hurdle to overcome in the initial phase of a pandemic, but can scale massively once they have been generated. Additionally, rapid tests only provide a binary readout (positive/negative), which can be difficult to interpret and the antigen is rarely specified (Sethuraman, Jeremiah, & Ryo, 2020). Due to their rapid turnaround and affordability, these tests can thus be deployed in millions to aid in large-scale screening efforts and by repeated testing over time, accuracy can be greatly improved (Mina, Parker, & Larremore, 2020; Ramdas, Darzi, & Jain, 2020).

In contrast to traditional PCR-tests or antigen rapid tests, LC coupled to Multiple Reaction Monitoring (MRM) tandem MS detection offers a straightforward assay toward pre-defined targets. Turning to MS measurements to detect SARS-CoV-2 in samples directly addresses the issue of specificity and the risk of returning false-positive results as the measurement benefits from the fundamental properties of MS detection of peptides through multiple specific product ions (Gillette & Carr, 2013), which results in sequence-based specificity through direct physical detection of analyte molecules. The instrumentation provides reliable quantification for absolute protein concentration determination and modern MS instrumentation offers unsurpassed specificity, high precision, excellent quantitative performance and high analytical sensitivity.

When combining these features with affinity reagents, such as antibodies, assays can reach very high sensitivities and low levels of a protein can be detected even in complex matrices. The combination of immuno-based strategies with mass spectrometry read-out can complement each other and provide target-specific protein quantification (Whiteaker & Paulovich, 2011). In fact, it is an ideal combination for rapid detection and reliable quantification of low abundance proteins. Stable isotope labeled (SIL) standards and capture by anti-peptide antibodies (SISCAPA) (N Leigh Anderson et al., 2004) enables multiplexed analysis of pre-digested clinical samples using peptide-reactive antibodies, selective for SARS-CoV-2 peptides, immobilized onto magnetic beads. Spiked SIL peptide standards further improve precise protein measurements performed by MRM (Brun, Masselon, Garin, & Dupuis, 2009). The use of LC-MS for protein quantification of SARS-CoV-2 peptides eliminates the dependence on PCR reactions and any issues related to unspecific amplification thanks to the selectivity achieved at three different levels: first by the antibody; secondly by the mass spectrometric read-out and; finally, the internal standard. As a proof of concept, we analyzed clinical samples collected from asymptomatic individuals screened for ongoing disease by RT-PCR. Samples were taken from the upper respiratory tract (combined triple-point collection strategy throat/nasopharynx/saliva) and a set of 48 PCR positive and 308 RT-PCR negative samples were selected for LC-MS analysis. All samples were analyzed using the SISCAPA immuno-affinity peptide enrichment protocol followed by LC-MS read-out outlined in **Figure 1**. The application of immuno-affinity peptide enrichment is typically associated with the detection of protein disease markers in body fluids, such as, plasma or dried blood spot samples.

**Figure 1.**
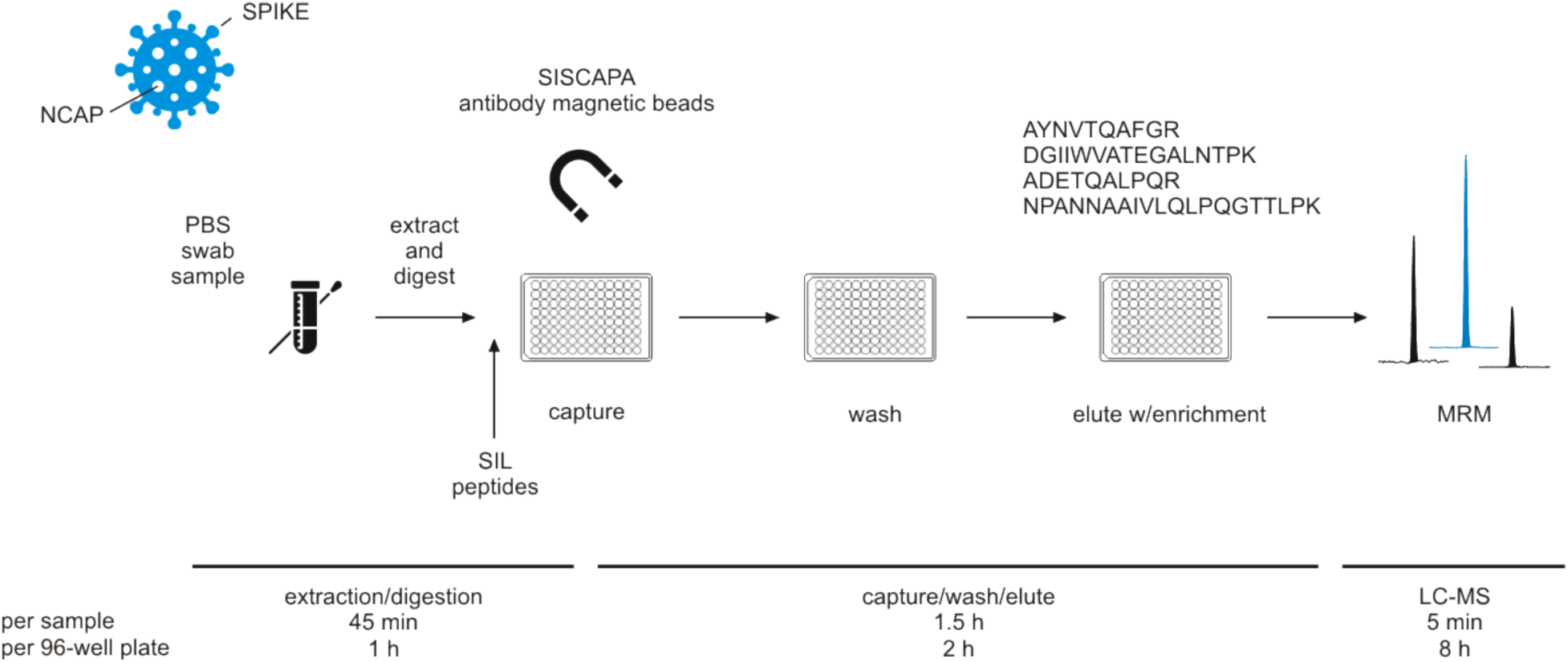
Experimental workflow for immuno-affinity peptide (SISCAPA) enrichment LC-MS of NCAP SARS-CoV-2 peptides. Swab sample extracts were subjected to tryptic digestion, SIL standards added to the tryptic digest solution, and magnetic beads coupled with specific anti-peptide antibodies incubated to allow binding of the peptides. Unbound peptides are removed and the target peptides eluted and measured using MRM analysis with LC-MS.

Here, the novel application of the technology is demonstrated to detect and quantify infection by analyzing the protein complement of viruses at relevant levels, which are proven difficult to reach without enrichment (Van Puyvelde et al., 2021). This study thereby presents a precise and complementary approach to RT-PCR to reliably detect SARS-CoV-2 in a research or clinical setting and a possible route forward to support population-wide screening.

## RESULTS

The application of LC-MS to detect tryptic digest peptides of SARS-CoV-2 proteins has been successfully demonstrated (Cardozo et al., 2020; Cazares et al., 2020; Freire-Paspuel & Garcia-Bereguiain, 2021; Gouveia, Grenga, et al., 2020a; Gouveia, Miotello, et al., 2020b; Ihling et al., 2020; Saadi et al., 2021; Van Puyvelde et al., 2021). However, these studies also highlight that the technique can be hampered by matrix effects, *i*.*e*., analysis interferences arising from the constituent components of swab (preservation) media or other matrices, as well as base sensitivity, to be able to reach clinically relevant detection levels, suggesting the need for clean-up, *e*.*g*., solid phase-based extraction, and/or affinity enrichment (Renuse et al., 2020; Van Puyvelde et al., 2021). Moreover, commonality can be observed within the results of these studies in terms of which tryptic digest peptides are typically detected by means of LC-MS. NCAP is the most abundant viral SARS-CoV-2 protein with an estimated ∼ 300 – 1,000 copies per virion particle (Bezstarosti, Lamers, van Kampen, Haagmans, & Demmers, 2020; Parks & Smith, 2020), making it, because of the relatively high number of NCAP copies per virion, an attractive target for LC-MS based detection compared to other viral proteins. A number of NCAP candidate peptides were therefore evaluated in terms of and LC-MS behavior, *i*.*e*., sensitivity and linear dynamic range, and peptide immunoassay suitability (Whiteaker et al., 2011). The LC-MS MRM responses of a number of candidate NCAP SIL peptides are shown in **Figure 2–figure supplement 1**, ranking the peptides in descending order of MRM sensitivity. From this set of peptides, primarily based on both MRM response and peptide immunoassay suitability, peptide AYNVTQAFGR was found to be one of the best surrogate peptide candidates, but, equally importantly, it is not significantly affected to date by known SARS-CoV-2 virus mutations (https://www.gisaid.org/). Other evaluated peptides, but not discussed in detail, included ADETQALPQR, DGIIWVATEGALNTPK and NPANNAAIVLQLPQGTTLPK, of which the basic quantitative characterization results are summarized in **Figure 2–figure supplement 2-4**, respectively.

### Method Characterization

The LC-MS MRM data were processed using TargetLynx XS and with a cut-off threshold algorithm based on peptide peak height and area thresholds, as well as quantifier to qualifier ion ratio threshold (30%). In other words, using two different consistently measured peptide fragment ions, *i*.*e*. MRM transitions, to confirm the presence of SARS-CoV-2 proteins. Typical detection examples for the quantifier, qualifier and SIL MRM transitions are shown in the (A) panel of **Figure 2**. An internal standard SIL corrected LC-MS calibration curve for antibody enriched NCAP peptide AYNVTQAFGR detected in a spiked nasopharyngeal swab matrix solution is shown in the (B) panel of **Figure 2**, covering a linear dynamic range from 3 to 50,000 amol/μl, providing > 4 orders of linear dynamic range, meanwhile affording an LLOQ amount of 3 amol/μl of AYNVTQAFGR peptide (with precision ≤20%, bias ±20% and S/N >10:1 (peak-to-peak)). Shown as well are example quantifier and qualifier MRM chromatograms of positive (**Figure 2C**) and negative (**Figure 2D**) SARS-CoV-2 PBS swab samples. The selectivity of the method is highlighted by the complete absence of signal in the MRM chromatogram of the negative SARS-CoV-2 sample (**Figure 2D**).

**Figure 2.**
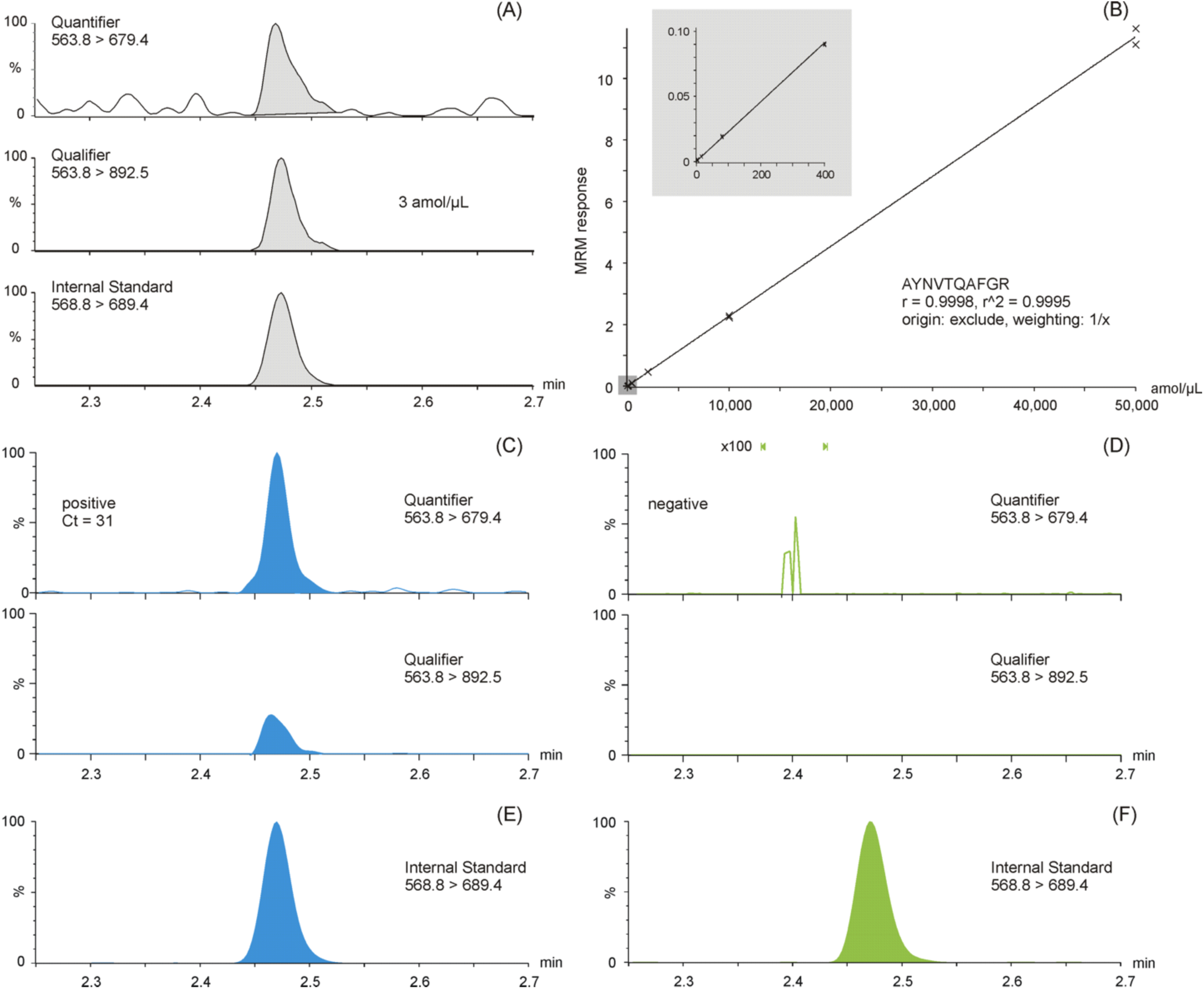
MRM chromatograms of antibody enriched NCAP AYNVTQAFGR peptide. Quantifier, Qualifier, and SIL Internal Standard peptide chromatograms spiked at the lower limit of quantification (3 amol/μl) **(A)**. Calibration curve of the AYNVTQAFGR peptide based on enriched recombinant NCAP protein digest, spiked with a constant amount of SIL peptide **(B)**. Two representative intensity-scaled MRM chromatograms of positive (mean Ct 31) **(C)** and negative (blank) **(D)** SARS-CoV-2 swab samples, respectively, normalized to the most abundant shared MRM transition. Intensity-scaled SIL Internal Standard peptide MRM chromatograms of positive **(E)** and negative **(F)** SARS-CoV-2 swab samples.

The precision of the method was evaluated at 3, 10, 400 and 25,000 amol/μl for NCAP AYNVTQAFGR peptide and NCAP protein spiked into PBS and viral transport medium (VTM, Liofilchem, Italy). Peptides were enriched by antibodies and samples were analyzed in replicates of 5-over-5 separate occasions. The inter- and intra-day precision values of the method, as summarized in **Table 1**, were shown to be ≤20 %CV.

**Table 1.** Intra- and inter-day method precision (n = 5) when monitoring SARS-CoV-2 NCAP peptide AYNVTQAFGR using immuno-affinity peptide enrichment LC-MS (MRM).

|  | precision (% CV) |  |  |  |  |  |  |  |
| --- | --- | --- | --- | --- | --- | --- | --- | --- |
|  | intra (concentration [amol/μL]) |  |  |  | inter (concentration [amol/μL]) |  |  |  |
|  | 3 | 10 | 400 | 25,000 | 3 | 10 | 400 | 25,000 |
| peptide-spiked PBS | 12.0 | 11.1 | 5.8 | 5.2 | - | - | - | - |
| NCAP-spiked PBS | 18.9 | 3.9 | 4.8 | 6.4 | - | - | - | - |
| peptide-spiked VTM | 12.5 | 6.8 | 2.4 | 3.0 | 15.5 | 10.2 | 6.8 | 4.7 |
| NCAP-spiked VTM | 13.2 | 10.2 | 2.4 | 2.9 | 11.6 | 17.6 | 18.5 | 11.1 |

- not tested

Additionally, the AYNVTQAFGR peptide was shown to be stable in the autosampler at 10°C for over 48 hours following re-analysis and comparison to a stored calibration curve.

### Sample Analysis

The samples analyzed by LC-MS and RT-PCR were compared. The High and Low Pools were analyzed in triplicate with a precision of 3.0% CV and 12.2% CV, respectively for each pool. Example quantifier and qualifier LC-MS MRM chromatograms of peptide AYNVTQAFGR are shown in the two bottom panes of **Figure 2**, respectively. The results shown in **Figure 3A** suggests good (inverse) correlation between the LC-MS (log_2_ transformed quantifier response, *i*.*e*, SIL corrected quantifier peak area) and the RT-PCR (Ct) data, which has also been noted in other so-called ‘non-enriched’ studies (Van Puyvelde et al., 2021). The results shown in **Figure 3B** represent the LC-MS data in an alternative, quartile distribution-based format, suggesting that differentiation between sample types is feasible and that the detected abundances are significantly different (p = 0.00018; Mann-Whitney U test).

**Figure 3.**
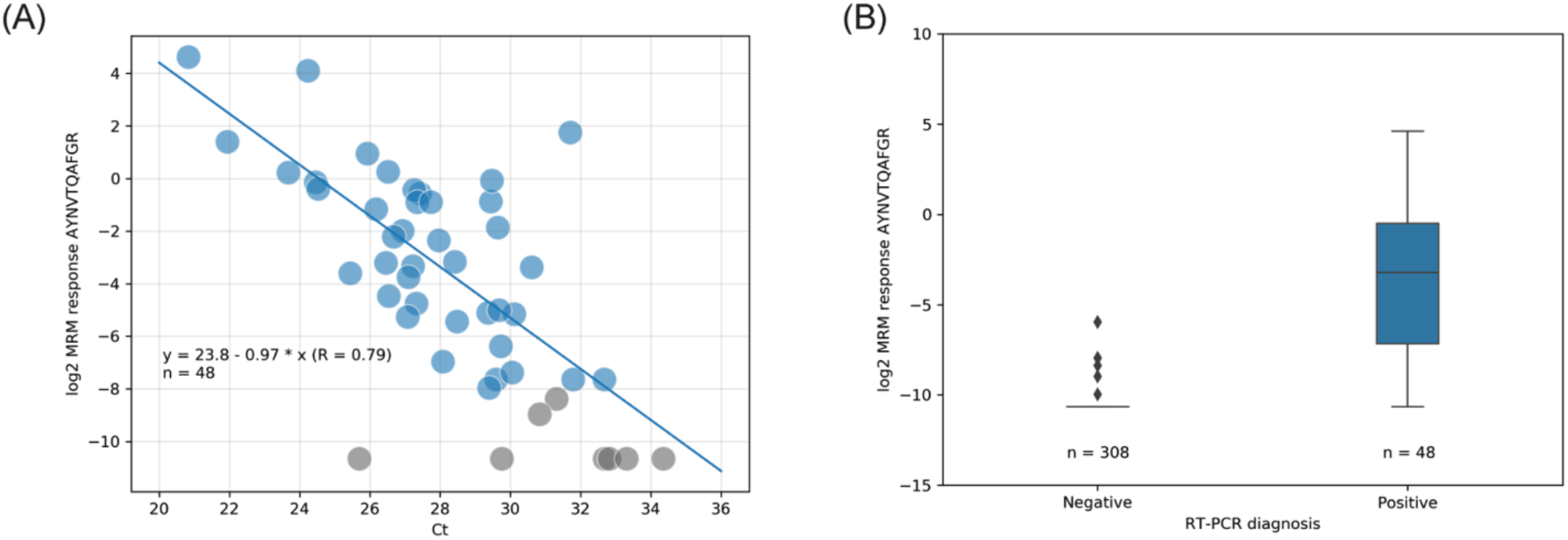
LC-MS (log_2_ quantifier response) vs. RT-PCR (Ct) read-out correlation with linear regression **(A)** and quartiles distribution of the LC-MS results **(B)**. Color labeling is based on RT-PCR diagnoses; blue = positive SARS-CoV-2; grey = not detected (no light signals) or inconclusively quantified (single transition) by LC-MS.

Following CLSI EP 12-A2 User Protocol for Evaluation of Qualitative Test Performance guidance, a summary of the sample analysis results is shown in a 2×2 contingency table format in **Figure 4**, using the RT-PCR results as a reference, estimated sensitivity and specificity values for LC-MS are 83.3% and 100%, respectively. The 95% score confidence interval limits for sensitivity calculations, were 70.4% to 91.3% and for specificity were 98.8% to 100%. Accordingly, the agreement between RT-PCR and LC-MS was strong (kappa value of 0.9 (95% CI 0.83 – 0.97). When analyzing samples above the estimated LLOQ (3 amol/μl, which approximates to Ct ≤30) the estimated sensitivity is improved to 94.7% with the corresponding 95% score confidence interval limits for sensitivity 82.7% to 98.5%. Further work will look at adding a secondary confirmatory peptide to the cut-off algorithm. However, RT-PCR does not distinguish between infectious virus and non-infectious nucleic acids (Engelmann et al., 2021), whereas LC-MS will only detect one or multiple peptides from the protein complement of the virus. This has implications on the interpretation of RT-PCR Ct levels itself in terms of infectious *vs*. non-infectious classification of patient samples but also for determining the sensitivity and specificity of complementary and/or alternative methods. Peptide levels have not been evaluated in the context of infectiousness yet, but other conditions, such as sample storage before LC-MRM/MS can also give rise to analytical variance due to the inherent difference in stability between RNA and proteins. Additionally, Ct values are not universally applicable as they differ between manufacturers and methods (Engelmann et al., 2021; van Kasteren et al., 2020), which enforces the need of methods that are capable of determining viral load more accurately.

**Figure 4.**
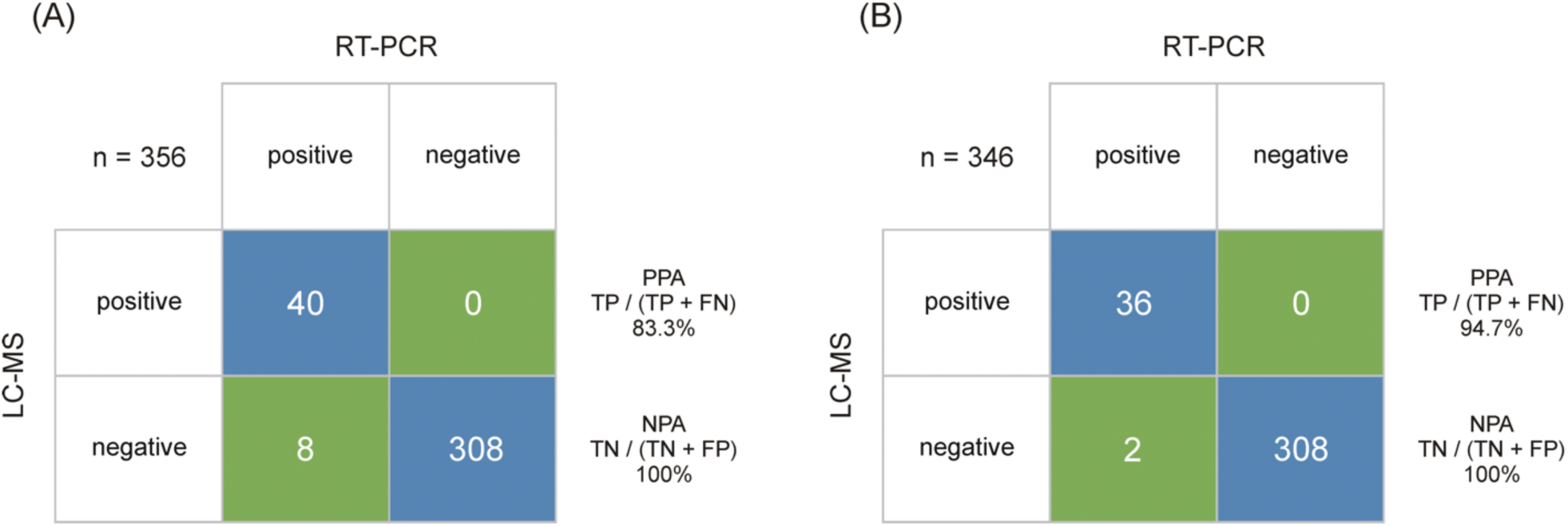
Output class (LC-MS) vs. target class (RT-PCR) contingency matrix, used to calculate the PPA and NPA of the SARS-CoV-2 immuno-affinity peptide enrichment LC-MS method (A) The LC-MRM/MS performance is based on RT-PCR results obtained from 48 positive and 308 negative samples. (B) The LC-MRM/MS performance based on all positive samples with a RT-PCR results below Ct 30 (LOD for the LC-MRM/MS) and 308 negative samples.

## DISCUSSION

Any diagnostic test result should be interpreted in the context of the probability of disease, but also include proper internal controls to ensure a high level of clinical specificity when used as a tool for large-scale screening. In this conceptual study, we have established an assay capable of detecting SARS-CoV-2 in self-collected samples. Multiple peptide assays were generated towards the NCAP protein and the assay that gave the best response was used to profile SARS-CoV-2 in clinical samples. The unsurpassed specificity of mass spectrometers combined with antibodies is an attractive route forward for a future molecular pandemic surveillance system. This specificity can help grow the assay repertoire, which can be expanded to cover multiple peptides or proteins by rather simple means, if anti-peptide antibodies are available. The SISCAPA peptide enrichment method ensures both high sensitivity and low risk of reporting false positives due to the combination of specific binding of the antibody (Hoofnagle & Wener, 2009) with LC-MRM/MS readout. This is achieved by multiple factors that greatly outperform RT-PCR and rapid antigen test in theory. First, antibodies are used to selectively enrich for the target peptide in a complex mixture. This helps increase the overall analytical sensitivity while LC-MS readily can distinguish between peptides in the separation and MRM steps. Secondly, internal standards added to the sample enable accurate and robust quantification. This provides an internal standard reference trace for every analyte and can help distinguish between false positive chromatographic peaks based on retention time and ion ratios, that in RT-PCR experiment would be reported as a positive due to the absence of internal standards and since each gene is detected by a single reporter dye.

The sensitivity can be further improved by increasing the sample load if needed. Additionally, the number of viral protein targets can also be scaled by introducing additional anti-peptide antibodies into the sample mixture. This would allow for an LC-MS based viral protein panel analysis method where relevant peptides, also including relevant spike-peptides for mutation surveillance, are monitored in an endemic scenario, either covering new emerging SARS-CoV-2 strains or other viruses, such as influenza or respiratory syncytial viruses.

We show that the SISCAPA technology is an attractive route forward for future molecular pandemic surveillance systems. The accuracy of the LC-MS-based method would tolerate low levels of positive samples without compromising the positive predictive value of large-scale screening efforts, and thereby providing a next-generation platform for disease surveillance and an attractive alternative to today’s RT-PCR based technologies.

## Material and Methods

### Sample collection

The study was performed in accordance with the declaration of Helsinki and the study protocol (“*Jämförande studier av Covid-19 smitta och antikroppssvar i olika grupper i samhället*”) was approved by the Ethical Review Board of Linköping, Sweden (Regionala etikprövningsnämnden, Linköping, DNR - 2020-06395). Informed consent and consent to publish, including consent to publish anonymized data, was obtained from all subjects. Briefly, asymptomatic individuals working at an elderly caregiver in Sweden were screened on a regular basis at their workplace. A three-point collection (throat, nasal, saliva) was performed by participants using a self-sampling collection kit (Sansure Biotech, Changsha, China) containing phosphate buffered saline (1x PBS, 137 mM NaCl; 2.7 mM KCl; 4.3 mM Na_2_HPO_4_; 1.47 mM KH_2_PO_4_). Clinical samples were collected by swabs dipped into the sample collection tube and transported to the laboratory within eight hours. All samples were heat inactivated upon arrival to ensure that the core temperature of the vial reached at least 56°C for 30 min. The protocol used ensured that the core temperature did not reach above 60°C ± 0.5°C (1sd), which has been shown to have no effect on the RT-PCR sensitivity.

### RT-PCR

Samples were analyzed using a RT-PCR test from Sansure Biotech (Changsha, China) according to FDA-EUA guidelines. The Novel Coronavirus (2019-nCoV) Nucleic Acid Diagnostic Kit was used for quantitative detection of the ORF-1ab and the N gene of novel coronavirus (2019-nCoV). Briefly, samples were lysed at room temperature for at least 10 minutes to allow for RNA release by chemical lysis using Sample Release Reagent (Sansure Biotech). The presence or absence of SARS-CoV-2 RNA was determined by RT-PCR combined with multiplexed fluorescent probing, which targets a SARS-CoV-2 specific region of ORF-1ab (FAM) and N gene (ROX) together with the human Rnase P internal control (Cy5). The RT-PCR analysis was performed using a CFX96 Real-Time PCR Detection System (Bio-Rad, Hercules, CA) programmed with the following RT-PCR protocol according to the manufacturer’s instruction [50°C, 30 min; 95°C 1 min] followed by 45 cycles of [95°C 30 s, 60°C 30 s]. The RT-PCR results were interpreted according to instructions. Positive [FAM/ROX Amplification, Ct <40]. Negative [FAM/ROX No amplification; Cy5 Amplification, Ct <40].

### Immuno-Affinity Peptide Enrichment LC-MS

#### Materials

Recombinant nucleocapsid protein (NCAP) was from R&D Systems, Minneapolis, MN, trypsin from Worthington, Lakewood, NJ, and anti-peptide antibodies from SISCAPA Assay Technologies, Washington DC. All other chemicals were from MilliporeSigma, St. Louis, MI, unless stated otherwise.

#### Calibrator preparation

NCAP digest, protocol described below, was used for calibration and quantitation of viral proteins. A serial dilution from 2.2 pmol/μl NCAP to 10,000, 2,000, 400, 80, 16, and 3 amol/μl was performed consecutively in pooled negative sample background.

#### Samples

Clinical samples subjected to two freeze-thaw cycles prior were anonymized and two control pools were established by pooling randomly chosen samples based on their RT-PCR result (Ct <30 [High Pool], 30≤Ct<33 [Low Pool]). A total of 180 μl from each sample was used per enrichment experiment. A set of 48 positive and 308 negative samples was subjected for the LC-MS analysis.

#### Protein extraction and digestion

A total of 20 μl of Denaturant Mixture (1 % (w/v) RapiGest (Waters Corporation, Milford, MA) in 1 M Triethylammonium bicarbonate (TEAB), 50 mM dithiothreitol (Waters Corporation) were aliquoted into the collection plate (Waters Corporation). Next, 180 μl of the diluted NCAP and patient samples were carefully transferred from the collection tubes into the same plate. The plate was incubated on a heater-shaker at 500 rpm at 56°C for 15 min followed by the addition of 50 μl trypsin solution (7.3 mg/mL trypsin in 10 mM HCl). After mixing at 500 rpm for 30 s. the samples were digested at 37°C for 30 min and thereafter quenched by addition of trypsin stopping agent (0.22 mg/mL of Tosyl-L-lysyl-chloromethane hydrochloride in 10 mM HCl) at a final concentration of 37 μg/mL. The sample plate was mixed at 500 rpm for 30 s and incubated at room temperature for 5 min. The samples were spiked with 20 μl of SIL peptide mixture solution and mixed thoroughly on a shaker at 500 rpm for 30 s.

#### Peptide Enrichment

Anti-peptide antibodies, raised towards proteotypic peptides from the NCAP protein, were screened and validated as previously described (Pope, Soste, Eyford, Anderson, & Pearson, 2009). The antibody-coupled magnetic bead immune adsorbents corresponding to four SIL peptides (ADETQALPQR-^13^C_6_^15^N_4_, AYNVTQAFGR-^13^C_6_^15^N_4_, DGIIWVATEGALNTPK-^13^C_6_^15^N_2_, and NPANNAAIVLQLPQGTTLPK-^13^C_6_^15^N_2_) were resuspended fully by vortex mixing. The suspension of each anti-peptide antibody tube was mixed together in 1:1 ratio and 40 μl of the mixture was added to each digest. The plate was mixed at 1400 rpm to ensure that beads were resuspended and thereafter incubated for one hour at 800 rpm at room temperature. After one hour incubation, the plate was placed on a magnet array (SISCAPA Assay Technologies). As soon as the beads had settled on the sides of each well (typically one min), the supernatant was removed. 150 μl of wash buffer (0.03% CHAPS, 1xPBS) was added to each sample and the beads were fully resuspending at 1400 rpm for 30 s and 450 rpm for another 30 s. The plate was placed on the magnet array again and the supernatant was removed. This step was repeated three times. The beads were subsequently resuspended in 50 μl elution buffer (0.5 % formic acid, 0.03% CHAPS) and incubated for 5 min at room temperature. The beads were discarded by transferring the eluent to a QuanRecovery plate (Waters Corporation) for LC-MS analysis.

#### LC-MS Detection and Quantification

Chromatography was performed on an ACQUITY UPLC I-Class FTN system, with Binary Solvent Manager and column heater (Waters Corporation). 20 μl of the enriched sample was injected onto a ACQUITY Premier Peptide BEH C18, 2.1 mm x 50 mm, 1.7 μm, 300 Å column (Waters Corporation) and separated using a gradient elution of mobile phase A containing laboratory LC-MS grade de-ionised water with 0.1% (v/v) formic acid, and mobile phase B containing LC-MS grade acetonitrile with 0.1% (v/v) formic acid. The gradient elution was performed at 0.6 mL/min with initial inlet conditions at 5% B, increasing to 28% B over 4.5 min, followed by a column wash at 90% B for 0.6 min and a return to initial conditions at 5% B. The total run time was 5.7 min, with a 6.5 min injection-to-injection cycle time.

A Xevo TQ-XS tandem MS (Waters Corporation, Wilmslow, UK) operating in positive electrospray ionization (ESI+) was used for the detection and quantification of the peptides. The instrument conditions were as follows: capillary voltage 0.5 kV, source temperature 150°C, desolvation temperature 600°C, cone gas flow 150 L/h, and desolvation gas flow 1000 L/h. The MS was calibrated at unit mass resolution for MS1 and MS2. Light and heavy labelled peptides were detected using MRM mode of acquisition with experimental details overviewed in **Table 2**.

**Table 2.**
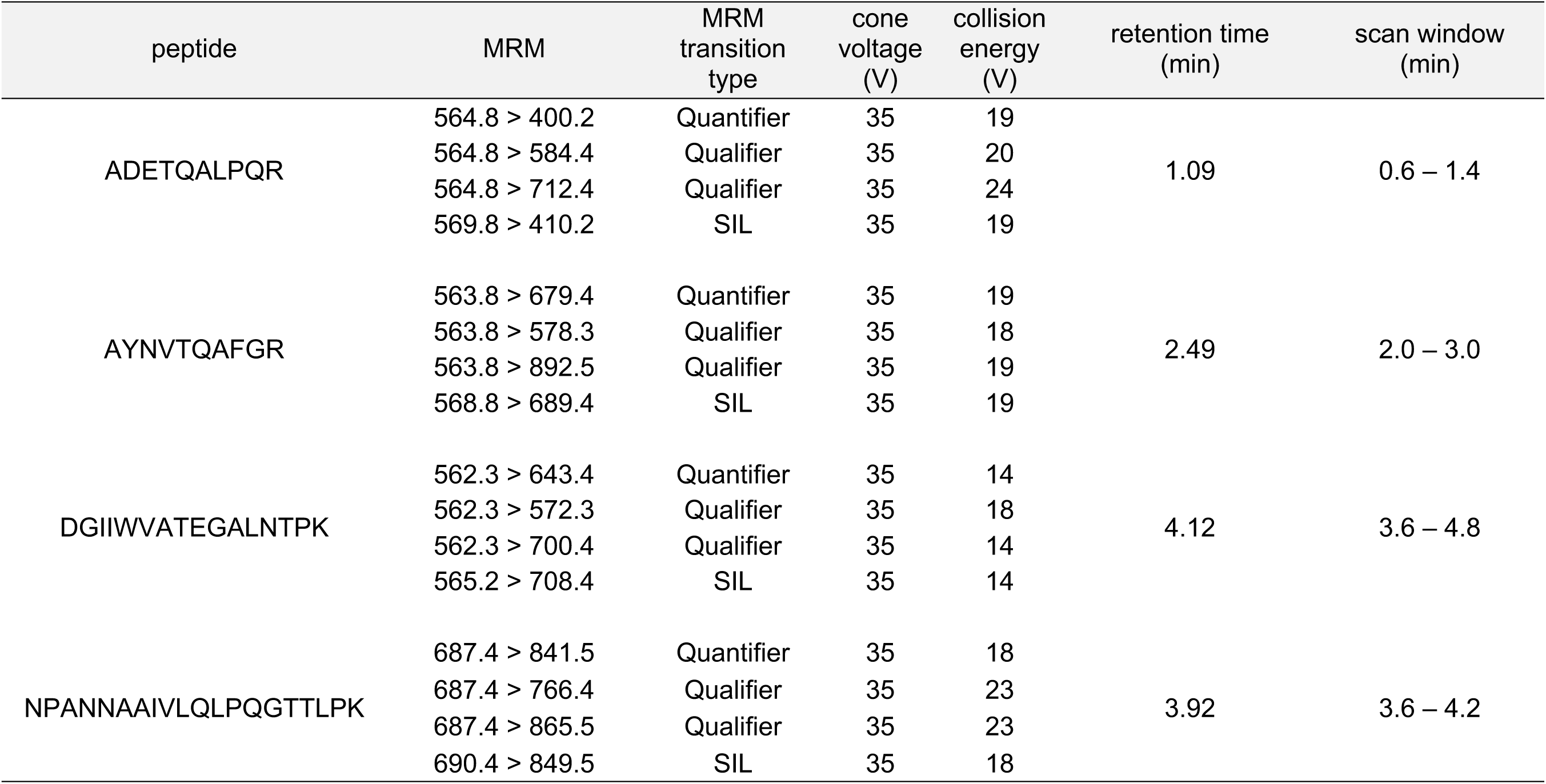
MRM transitions and MS method details target NCAP SARS-CoV-2 peptides.

TargetLynx XS (Waters Corporation) was used to process the raw LC-MS data, *i*.*e*., signal processing (mean smoothing and background subtraction), peak detection (area and height) and quantification of the MRM chromatograms, including the calculation of the quantifier ion to qualifier ion ratio. The quantified data were exported as tables (**Supplementary File 1**) and additional analysis and visualization carried out using Python 3.

## Data Availability

The proteomics data have been deposited to Panorama Public [38] (https://panoramaweb.org/sars-cov-2_siscapa.url; login, pwd: scilifelab), allowing for access to raw files and integrated peak areas from as well as visualization of LC-MRM/MS chromatograms.

## Data and materials availability

The ProteomeXchange ID for this dataset is PXD026366. The proteomics data have been deposited to Panorama Public (Sharma et al., 2014) (https://panoramaweb.org/sars-cov-2_siscapa.url; Password: LauwPWgd), allowing for access to raw files and integrated peak areas from as well as visualization of all LC-MRM/MS chromatograms.

## List of Supplementary Figures

**Figure 2 - figure supplement 1**. Peak area (MRM sensitivity) SIL (^13^C_6_^15^N_2_ C-terminal K or ^13^C_6_^15^N_4_ C--terminal R labeled) NCAP peptides as function of peptide and detergent (CHAPS) concentration.

**Figure 2 – figure supplement 2**. Calibration curve for ADETQALPQR over the range 3-50,000 amol/μl

**Figure 2 – figure supplement 3**. Calibration curve for NPANNAAIVLQLPQGTTLPK over the range 3-50,000 amol/μl

**Figure 2 – figure supplement 4**. Calibration curve for DGIIWVATEGALNTPK over the range 3- 2,000 amol/μl

